# Reproducibility of different methods of measuring left ventricular ejection fraction in echocardiography

**DOI:** 10.1101/2024.10.10.24315216

**Authors:** Ikuko Kawagishi, Masaya Hashimoto, Yuki Endo, Soma Ono, Miyabi Takamuku, Kazunori Ohnishi

**Affiliations:** Department of Clinical Laboratory, Faculty of Medical Science, Shubun University, Aichi, Japan

**Keywords:** echocardiography, reproducibility, left ventricular ejection fraction (LVEF), Teichholz method, modified Simpson method

## Abstract

**Background:** Left ventricular ejection fraction (LVEF) measurements by echocardiography are subject to problems such as low interobserver reproducibility, which depends on the skill of the observer, and variations in measurement technique.

**Objective:** We investigated the reproducibility of echocardiography by comparing the measurement results of a certified sonographer with those of control students.

**Methods:** A total of 30 subjects, 22 adult students and 8 staff members of Shubun University, were included in the study. Measurements were performed according to the standard protocols of the Japanese Society of Echocardiography.

**Results:** LVEF values were significantly lower with the modified Simpson method than with the Teichholz method (M-mode and B-mode) for both the sonographer and students; there was no difference between the M-mode and B-mode approaches for the sonographer, but B-mode was significantly lower for students. Comparing the sonographer and students, there was no significant difference with the M-mode approach, but students had significantly lower values with the B-mode and modified Simpson methods. Although the modified Simpson method is currently recommended for LVEF measurement, the lower values obtained by students using this method are primarily attributed to their inexperience with the measurement technique.

**Conclusions:** In the future, the evaluation of cardiac function by echocardiography will require reproducibility that does not rely on the skill of the observer, and the adoption of echocardiography methods incorporating artificial intelligence is anticipated.

## Introduction

Cardiac function comprises cardiac contractility (contraction force and rate) and diastolic function, with left ventricular ejection fraction (LVEF) being the most commonly used measure of cardiac function. Cardiac output is expressed as the product of stroke volume and heart rate. Various methods have been developed to measure this cardiac output, including echocardiography (2D and 3D), nuclear medicine imaging, CT imaging and cardiac magnetic resonance imaging (CMR). Accurate measurement of true cardiac output requires precise determination of left ventricular end-diastolic and end-systolic volumes, with CT or MRI being closest to true values. However, echocardiography is clinically practical due to its convenience, speed, and cost-effectiveness, though improvements are needed in reproducibility.

In this study, we measured LVEF using various methods, primarily contrasting young students with a certified sonographer. Both students and the sonographer performed measurements simultaneously. We compared measurements between different operators and within the same operators, evaluating accuracy, reproducibility, and utility. Additionally, we simultaneously measured the E-point septal separation (EPSS), a method used in emergency settings, and examined the pitfalls for beginners, such as students, to master the techniques.

## Subjects and methods

### Subjects

The study involved 30 healthy adult individuals from Shubun University, including 22 students and 8 staff members. The participants consisted of 28 males and 2 females, with an average age of 28.9 years (median 21 years; range 20 to 69 years). This study was conducted with the approval of the Research Ethics Committee of Shubun University Faculty of Medical Science (approval number: Medical Sciences R06-001).

### Ultrasonographic device and probe selection

The equipment used was the Fukuda Denshi Sefius UF-890 AG. The measurements were conducted by four students from Shubun University Faculty of Medical Sciences and one JSUM (the Japan Society of Ultrasonic in Medicine) registered sonographer (hereafter referred to as “the sonographer”). Prior to the study, each student who had completed a lecture and practical training in echocardiography furthermore received by six special training sessions.

### Measurement methods and sites

The following parameters were measured: aortic diameter, left atrial diameter, left ventricular septal thickness, posterior left ventricular wall thickness, left ventricular end-diastolic diameter, left ventricular end-systolic diameter, and left ventricular internal diameter shortening fraction. These were assessed using the Teichholz method, M-mode and B-mode. For measuring left ventricular ejection fraction (LVEF), the Teichholz method (M-mode and B-mode) and the modified Simpson method (biplane: Bp, 2-chamber: 2C, 4-chamber: 4C) were employed. The modified Simpson method involved calculating left ventricular end-systolic and end-diastolic volumes from apical 4-chamber and 2-chamber views. In tracing the ventricular cavity using the modified Simpson method, the papillary muscles, trabecular layer, and chordae tendineae were included within the ventricular cavity. Statistical analysis was performed using the EZR software (version R4.3.1). The correlation with body surface area (BSA) was assessed using Pearson’s correlation coefficient, while comparisons between students and the sonographer were tested using paired t-tests.

## Results

### 1. Echocardiographic parameters and correlations with individual echocardiographic measurements and body surface area (BSA)

Comparing the results of the present measurements with the JAMP study, which showed normal values of LVEF in a large number of healthy Japanese, LVEF was almost the identical in the sonographer, but lower in students.^1^ Table 1 shows each measurement separately for the sonographer and students. When comparing the measurements necessary for calculating LVEF using M-mode and B-mode between the sonographer and students, the left ventricular end-diastolic diameter was significantly lower in students: the sonographer 50.3 mm, while students 46.0 mm (p<0.0001). Similarly, the left ventricular end-systolic diameter was also significantly lower in students: sonographer 31.3 mm, while students 29.5 mm (p<0.012). However, there was no significant difference in left ventricular diameter shortening fraction, with the sonographer at 37.9% and students at 36.9%.

**Table 1.**
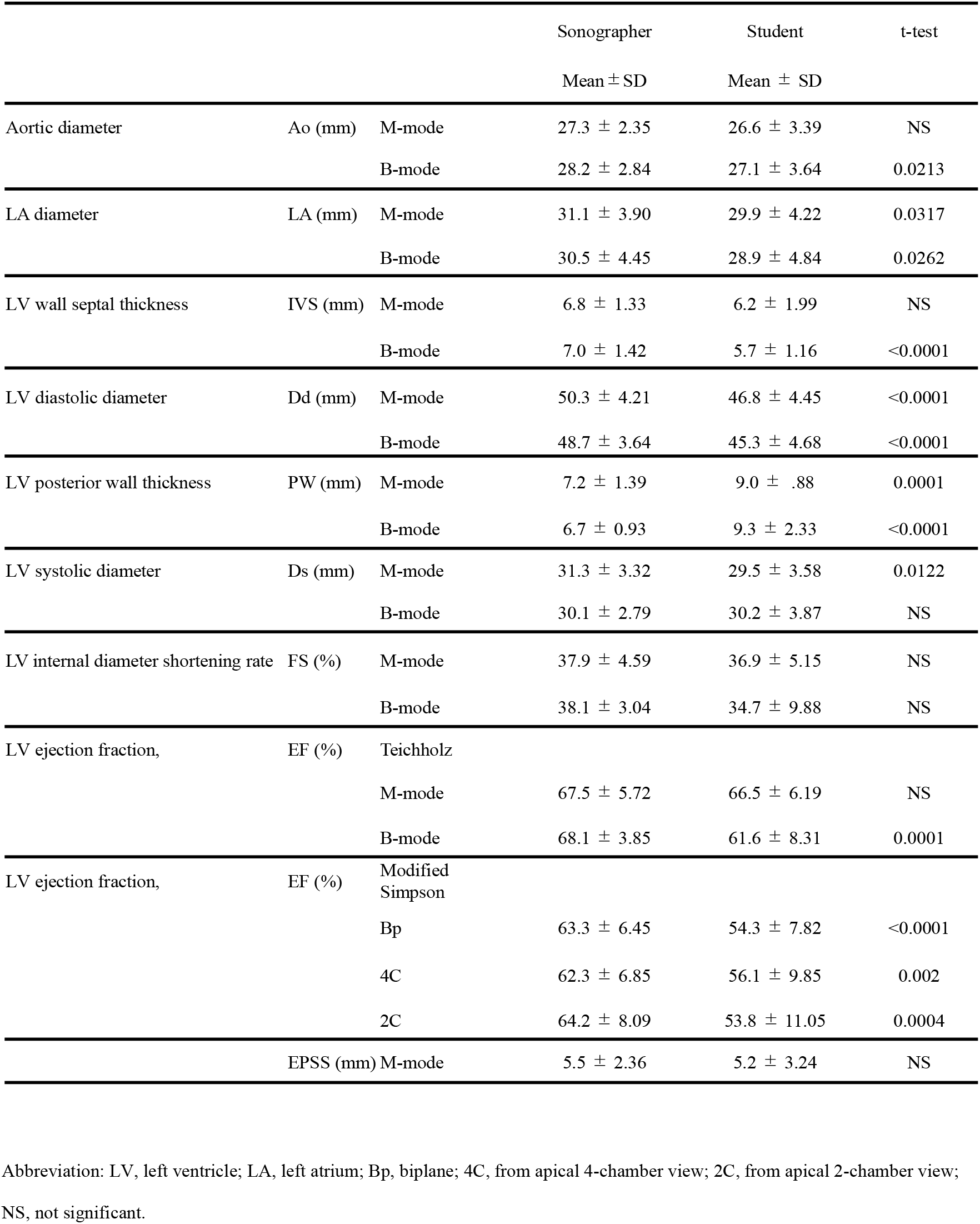
Echocardiographic measurements in this study.

When comparing left ventricular end-diastolic diameter, end-systolic diameter, and diameter shortening fraction using echocardiography between the sonographer and students, the left ventricular end-diastolic diameter was significantly lower in students: the sonographer 48.3 mm, while students 45.3 mm (p<0.0001). However, there was no significant difference in left ventricular end-systolic diameter, with the sonographer 30.1 mm and students 30.2 mm. Additionally, there was no significant difference in left ventricular diameter shortening fraction, with the sonographer at 38.1% and students at 34.7%.

Measurements of various cardiac parameters using M-mode and B-mode did not always show higher values with the M-mode. For M-mode measurements, the correlation between left ventricular end-diastolic diameter and BSA showed an increasing trend for both the sonographer and students, but it was not statistically significant (p = 0.079, p = 0.11) (Figure 1). The correlation between left ventricular end-systolic diameter and BSA was not significant for either group. The correlation between LVEF and BSA was not significant for the sonographer and students for both M-mode, B-mode and the modified Simpson method.

**Figure 1.**
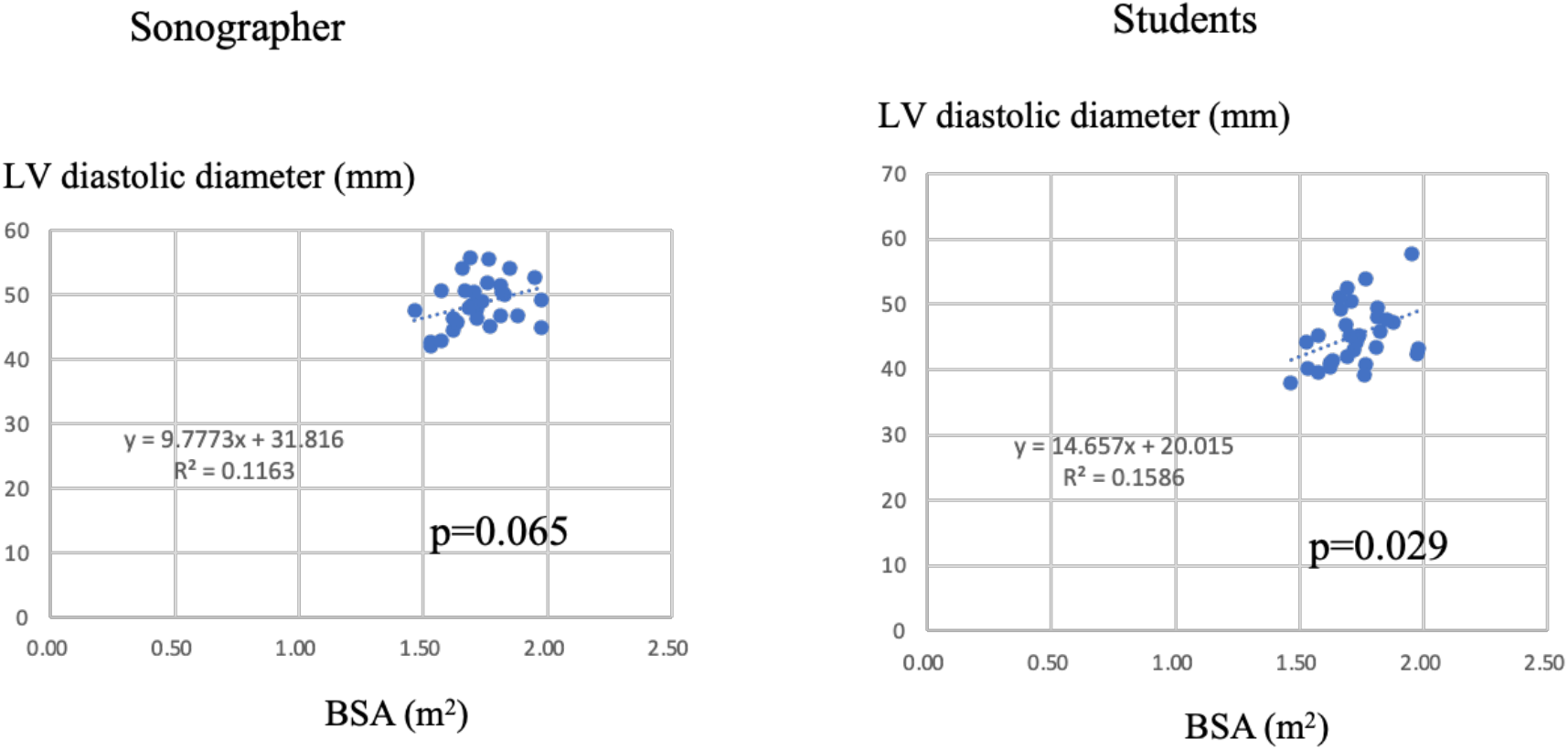
Correlation between left ventricular end-diastolic diameter and BSA

### 2. Comparison and correlation of LVEF measurements by the same operator

For both the sonographer and students, the average LVEF values measured using the modified Simpson method were significantly lower compared to those obtained using the Teichholz method (p < 0.001) (Figures 2,3). Specifically, the LVEF (mean ± SD) values were as follows: for the sonographer, M-mode method: 67.5 ± 5.7%, B-mode method: 68.1 ± 3.9%, modified Simpson method: 62.3 ± 6.4% (p < 0.0001) (Figure 2), and for students, M-mode method: 66.5 ± 6.2%, B-mode method: 61.6 ± 8.3%, modified Simpson method: 54.3 ± 7.8% (p < 0.01) (Figure 3).

**Figure 2.**
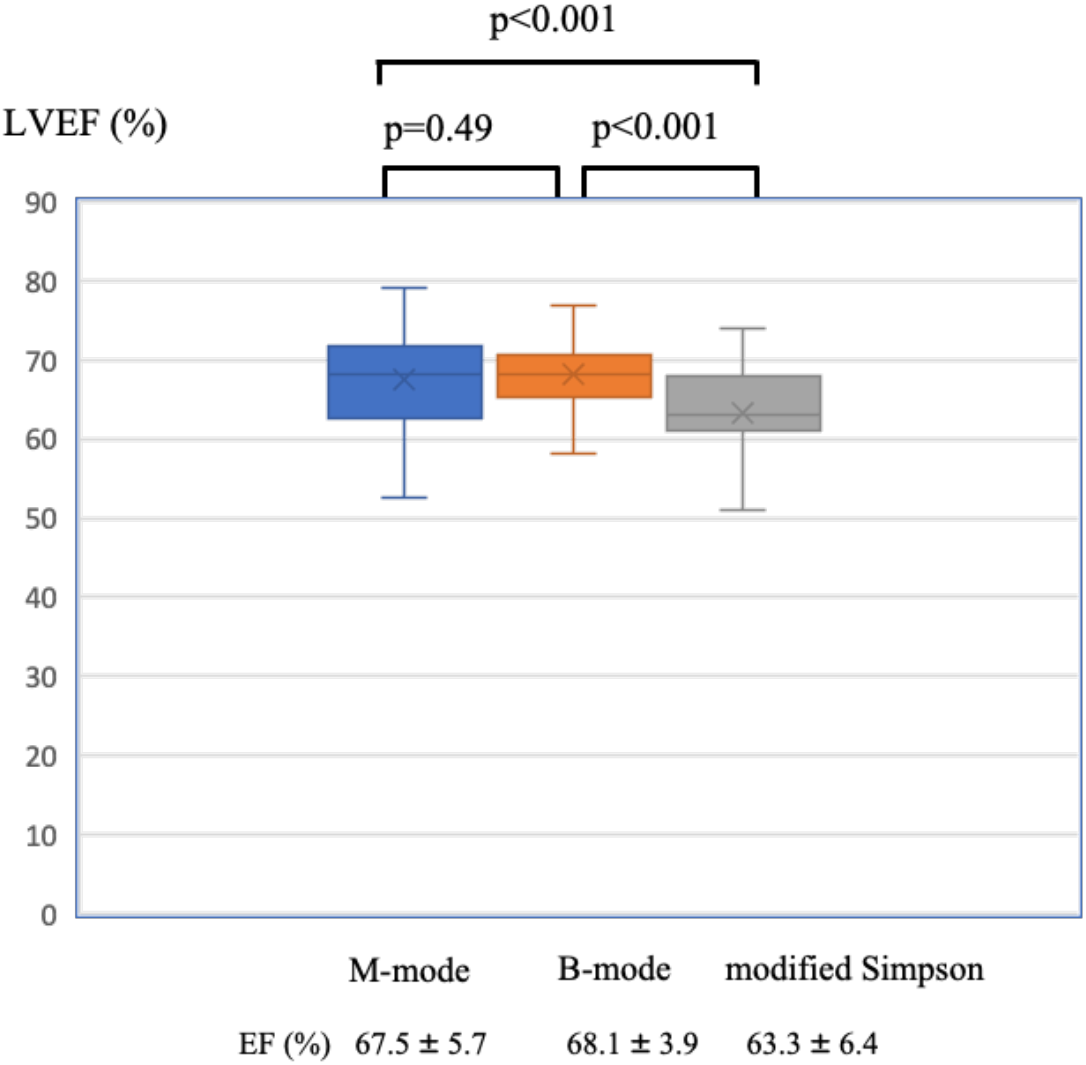
Comparison of LVEF between the Teichholz and the modified Simpson methods in the certified sonographer

**Figure 3.**
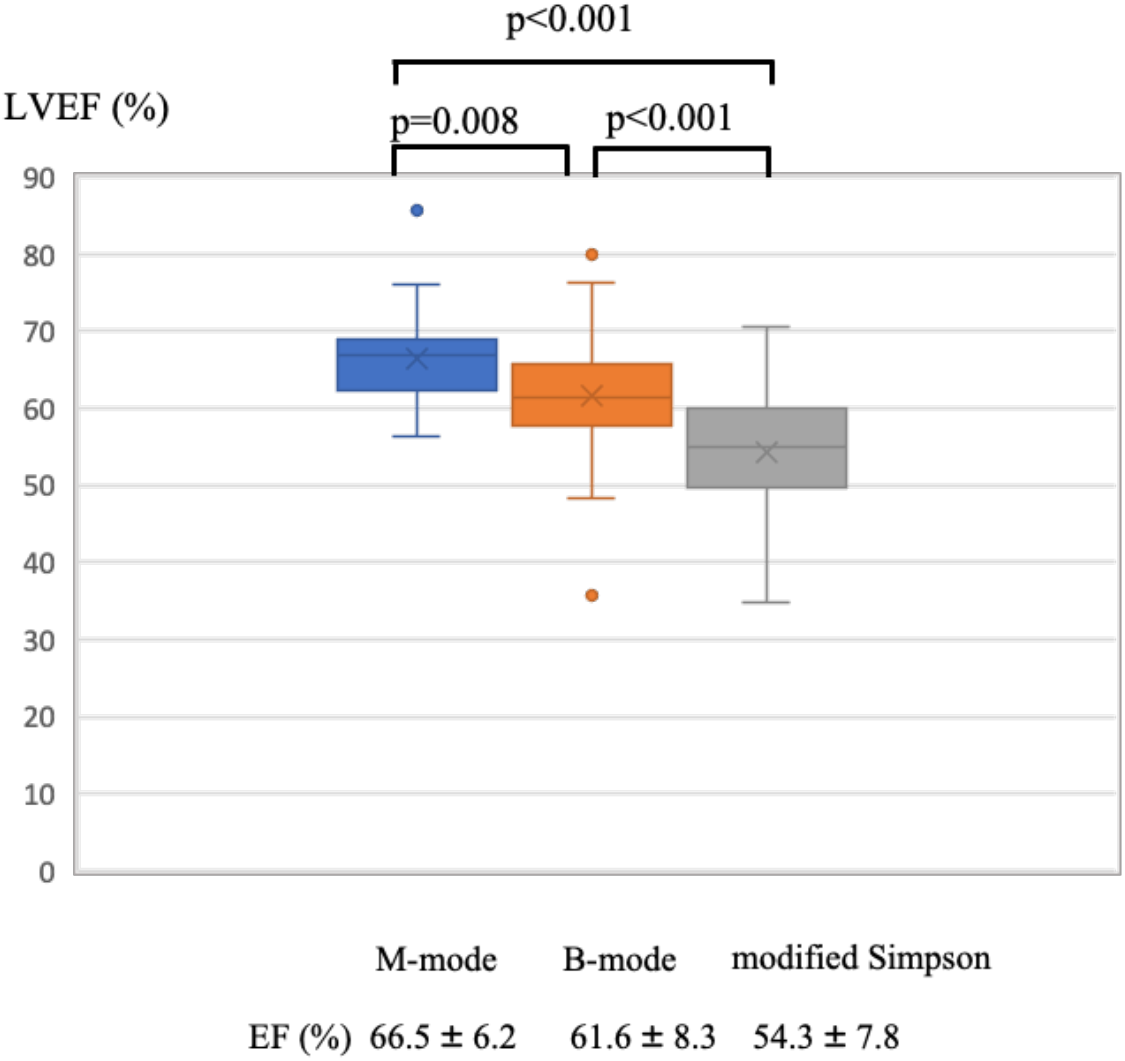
Comparison of LVEF between the Teichholz and the modified Simpson methods in students

Additionally, for both the sonographer and students, there was a significant correlation between M-mode and B-mode methods in LVEF measurements using the Teichholz method (p = 0.00029). However, no significant correlation was observed between the Teichholz method and the modified Simpson method, or between M-mode and Simpson method, or B-mode method and modified Simpson method (p = 0.18, p = 0.12) (Figure 4).

**Figure 4.**
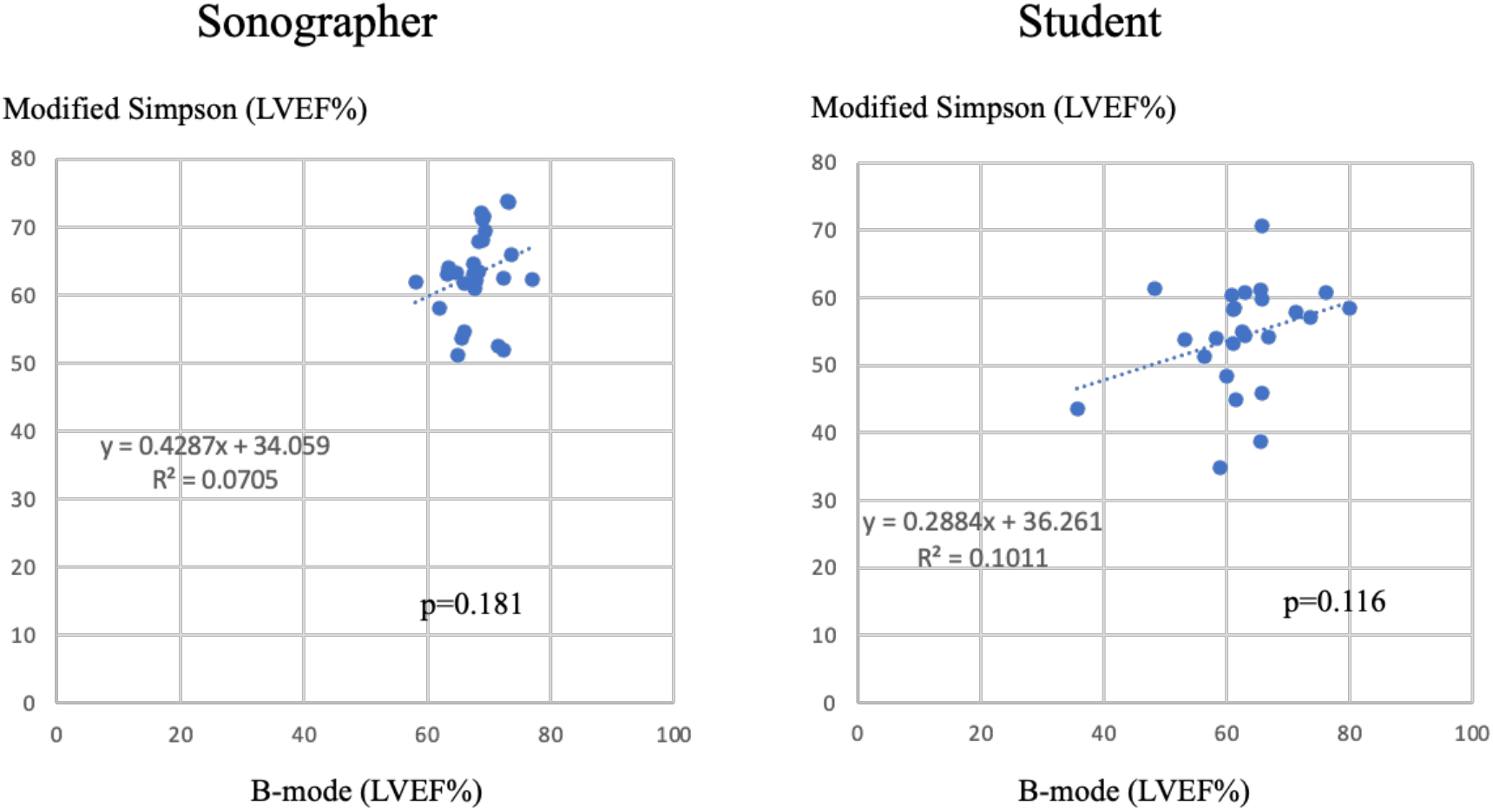
Correlation of the B-mode and the modified Simpson methods for LVEF in the sonographer and students

### 3. Comparison between the sonographer and students in measuring LVEF

LVEF (mean ± SD) was not significantly different between the sonographer and students in the M-mode method: the sonographer: 67.5 ± 5.7%, students: 66.5 ± 6.2% (p=0.46), while students had significant lower value in the B-mode method: the sonographer: 68.1 ± 3.8%, students: 61.6 ± 8.3% (p<0.001). In the modified Simpson method, LVEF was significantly lower in students: the sonographer: 63.3 ± 6.4%, students: 54.3 ± 7.8% (p<0.0001) (Figure 5). The M-mode method showed the highest reproducibility when comparing the sonographer and students, while the modified Simpson method showed the largest discrepancy.

**Figure 5.**
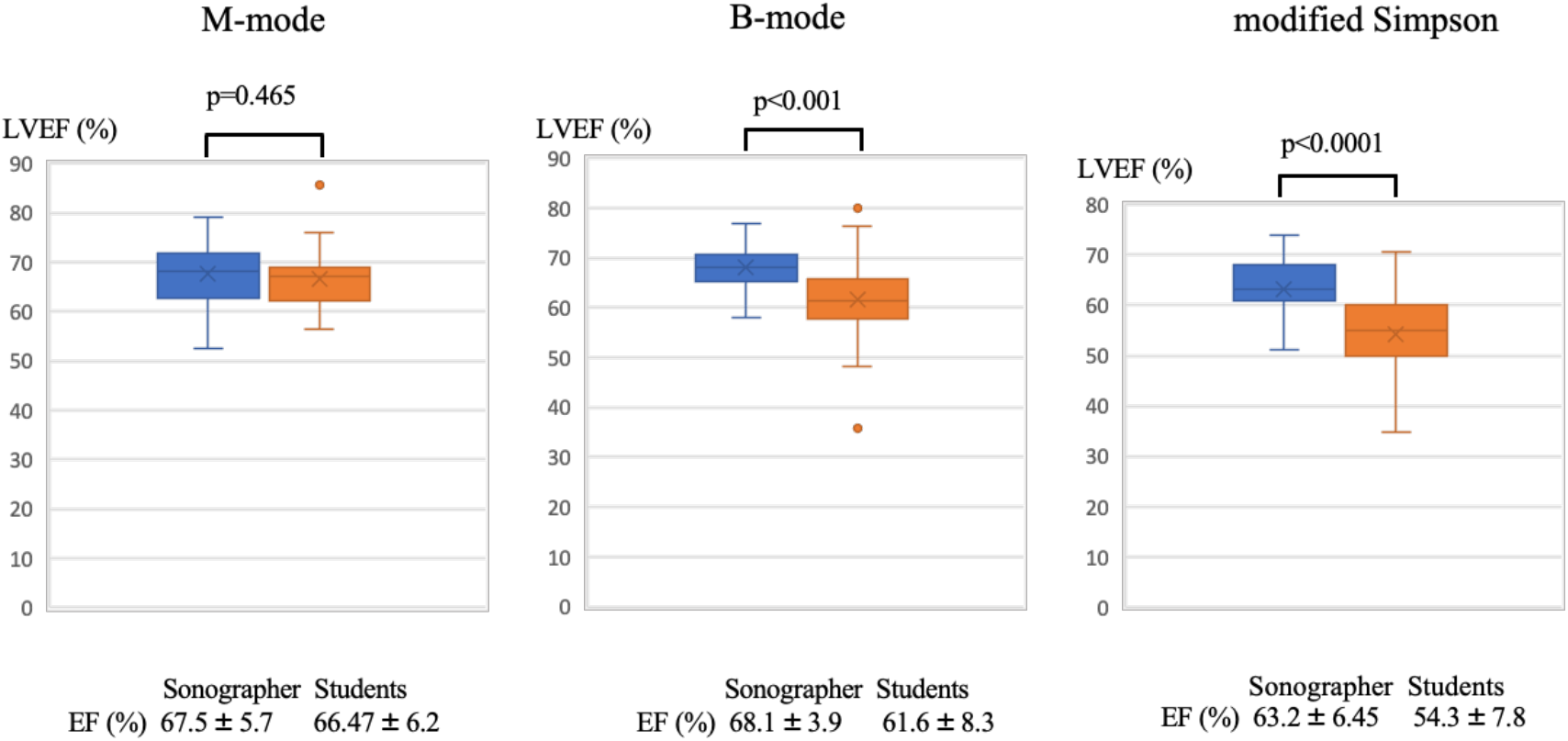
Comparison of LVEF measurements for the same subjects by the sonographer and students

The EPSS measurements were 5.53 ± 2.4 mm for the sonographer and 5.16 ± 3.24 mm for students (p=0.54), with no significant differences between them (Fig. 6).

**Figure 6.**
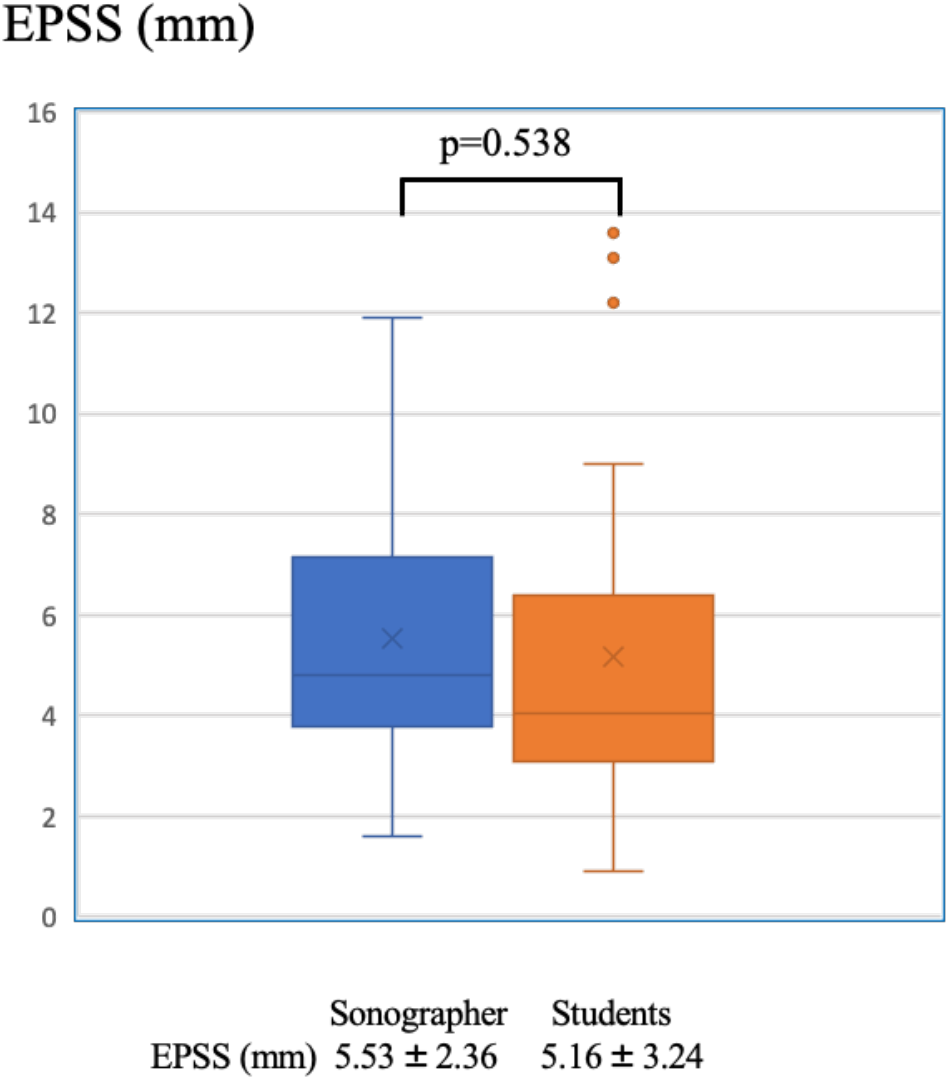
Comparison of EPSS measurements on the same subject by the sonographer and students

### 4. Correlation between EPSS and LVEF by the Teichholz and the modified Simpson methods

The correlation between EPSS and LVEF was significant for the M-mode method and B-mode method. Specifically, for the sonographer, there was a significant correlation between EPSS and LVEF measured using the M-mode method (p = 0.031), and for students, there was a significant correlation with the B-mode method (p = 0.012). However, no significant correlation was found between EPSS and LVEF measured by the modified Simpson method (the sonographer : p = 0.61, students: p = 0.56).

## Discussion

The measurement of LVEF using echocardiography has significant limitations, including poor interobserver reproducibility dependent on the observer’s skill, poor acoustic windows, and variability in measurement techniques.^2^ In the present study, we focused on the variability of measurement results between the certified sonographer and beginners.

Our findings confirm that LVEF values vary depending on the measurement method. The Teichholz method yielded significantly higher values compared to the modified Simpson method for both the certified sonographer and students. When comparing the M-mode and the B-mode methods in the Teichholz method, there was no significant difference for the sonographer, but students showed significantly lower values with the B-mode method. Additionally, while there was no significant difference in LVEF measurements using the M-mode method between the sonographer and students, students had significantly lower LVEF values with the B-mode and the modified Simpson methods. Thus, the M-mode method demonstrated the highest reproducibility when comparing the sonographer and students. The B-mode method aims to minimize interobserver error and is recommended in current guidelines for left ventricular diameter measurements in echocardiography.^3,4^ However, the variability in LVEF values with the B-mode method was greater among students compared to the M-mode method. This is likely due to fewer oblique measurements in the young, healthy population studied.

The modified Simpson method showed the largest discrepancy between the sonographer and students, which was mainly due to difficulties in inadequate apex delineation and inaccurate tracing of the left ventricular lumen. No significant differences were found in EPSS measurements. Normal LVEF values for healthy Japanese adults have been reported as 64 ± 5% for males and 66 ± 5% for females, and the measurements obtained from the sonographer for students and staff at our university were consistent with these values.^1^ The following discussion focuses on the reproducibility of echocardiography.

In echocardiography, the Teichholz method has traditionally been used to estimate left ventricular volume by assuming that the heart is a rotating ellipsoid and calculating the volume from left ventricular end-diastolic and end-systolic diameters. However, its accuracy has been questioned as it may not be suitable in cases where the left ventricle is flattened or spherically shaped. Currently, the most recommended method for measuring left ventricular volume is the biplane disc summation method (the modified Simpson method) using 2D echocardiography.^3,4^ This method calculates LVEF by summing the volumes of 20 discs approximating the left ventricular volume. The left ventricular volume is derived from apical 4-chamber and 2-chamber views to compute LVEF. Recently, the left ventricular longitudinal axial systolic function assessment (global longitudinal strain) has been shown to detect reduced systolic function earlier than LVEF and to be useful in predicting prognosis in patients with cardiac disease.^3^

On the other hand, cardiac magnetic resonance (CMR) measurements are currently considered the most accurate method of measuring cardiac function.^5^ The reproducibility of CMR measurements is superior to echocardiography in most studies. Anatomically, the structure of the left ventricular wall consists of a columnar layer and a dense layer, with the tip of the columnar layer being endocardial. Echocardiography measures volume by tracing the columnar layer (papillary muscle, columnar layer and tendon cords are included in the cardiac cavity), while CT and CMR measure volume by tracing the dense layer, which difference can introduce discrepancies. In other words, left ventricular volume measured by echocardiography is underestimated compared to CT and CMR.^3^ A 2014 report also indicated that contrast-enhanced 2D echocardiography and non-contrast 3D echocardiography showed good reproducibility and good agreement with LVEF measured by CMR, although volume concordance was reported to be poor.^2^ Dorosz et al. reported that left ventricular volume measured by 2-dimentional (2D) echocardiography was underestimated by an average of 48.2 mL when compared to that measured by CMR.^6^ Furthermore, the modified Simpson variant has a measurement error of approximately 10% in LVEF.^7,8^

In the field of anticancer treatment-related cardiac dysfunction, this LVEF value is also used as a diagnostic criterion and more reproducible methods are required. Recently, the definition of anticancer treatment-related myocardial impairment is often defined as ‘LVEF below 53% with a 10 % reduction from baseline’.^9^ However, depending on the measurement method, 10% may be within the margin of error and is considered problematic.

The modified Simpson method in echocardiography exhibits significant variability depending on the section imaged. To improve measurement accuracy, it is essential to capture the true apical 4-chamber view and keep the difference in left ventricular long-axis measurements between the two views within 10%. The American Society of Echocardiography guideline recommends using 3D echocardiography for left ventricular volume measurement if the true apical view cannot be obtained.^4^ The significant differences observed in the measurements by students compared to the sonographer are likely due to difficulties in capturing the true apical view and inaccuracies in tracing the left ventricular cavity.

Recently, the results of a blinded, randomized trial of cardiac function assessment in echocardiography by the sonographers and artificial intelligence (AI) were reported.^10^ The agreement rates between the measurements by the sonographers or AI and the final evaluations by cardiologists showed no significant differences. An AI-guided workflow for the initial assessment of cardiac function in echocardiography was non-inferior and even superior to the initial assessment by the sonographers.

On the other hand, the American College of Intensive Care Medicine guideline recommends a rapid qualitative assessment of left ventricular function in intensive care environments. Qualitative estimates of left ventricular systolic function using visual assessment (eye ball EF) by experienced sonographers have been shown to be accurate in several studies. However, the disadvantages of this method are the need for training and observer dependence. Additionally, EPSS (E-point septal separation) has been studied as a tool for emergency physicians to evaluate LVEF at the bedside. EPSS measurements (<5 mm is considered normal) have been shown to correlate strongly with LVEF by the Teichholz method.^12^ In the future, there will be a need for more rapid and reproducible quantitative evaluations of LVEF, including in emergency settings, with expectations for increased utilization of AI.

In conclusion, in this comparison between the sonographer and students in the measurement of LVEF by echocardiography, the M-mode method had the best reproducibility, while the modified Simpson method showed the largest discrepancy. The low LVEF values in the modified Simpson in students were considered to be mainly due to inexperience with the measurement technique. Echocardiography will continue to be the predominant method of assessing cardiac function from the perspective of simplicity, speed and cost-effectiveness. As the clinical indications for echocardiography increase, it is essential that these measurements are used reliably for accurate diagnosis and serial assessment of cardiac function. Therefore, AI-guided echocardiography and other techniques are likely to become more important in the future.

Disclosure of Conflicts of Interest (COI): The authors report no conflicts of interest.

## Data Availability

All data produced in the present study are available upon reasonable request to the authors

